# Aerosol and surface stability of HCoV-19 (SARS-CoV-2) compared to SARS-CoV-1

**DOI:** 10.1101/2020.03.09.20033217

**Authors:** Neeltje van Doremalen, Trenton Bushmaker, Dylan H. Morris, Myndi G. Holbrook, Amandine Gamble, Brandi N. Williamson, Azaibi Tamin, Jennifer L. Harcourt, Natalie J. Thornburg, Susan I. Gerber, James O. Lloyd-Smith, Emmie de Wit, Vincent J. Munster

**Affiliations:** Laboratory of Virology, Division of Intramural Research, National Institute of Allergy and Infectious Diseases, National Institutes of Health, Hamilton, MT, USA; Department of Ecology and Evolutionary Biology, Princeton University, Princeton, NJ, USA; Department of Ecology and Evolutionary Biology, University of California, Los Angeles, Los Angeles, CA, USA; Division of Viral Diseases, National Center for Immunization and Respiratory Diseases, Centers for Disease Control and Prevention, Atlanta, GA, USA; Fogarty International Center, National Institutes of Health, Bethesda, MD, USA

## Abstract

A novel human coronavirus, now named severe acute respiratory syndrome coronavirus 2 (SARS-CoV-2, referred to as HCoV-19 here) that emerged in Wuhan, China in late 2019 is now causing a pandemic^1^. Here, we analyze the aerosol and surface stability of HCoV-19 and compare it with SARS-CoV-1, the most closely related human coronavirus.^2^ We evaluated the stability of HCoV-19 and SARS-CoV-1 in aerosols and on different surfaces and estimated their decay rates using a Bayesian regression model (see Supplementary Appendix). All experimental measurements are reported as mean across 3 replicates.

---

HCoV-19 remained viable in aerosols throughout the duration of our experiment (3 hours) with a reduction in infectious titer from 10^3.5^ to 10^2.7^ TCID_50_/L, similar to the reduction observed for SARS-CoV-1, from 10^4.3^ to 10^3.5^ TCID_50_/mL (Figure 1A). HCoV-19 was most stable on plastic and stainless steel and viable virus could be detected up to 72 hours post application (Figure 1A), though the virus titer was greatly reduced (plastic from 10^3.7^ to 10^0.6^ TCID_50_/mL after 72 hours, stainless steel from 10^3.7^ to 10^0.6^ TCID_50_/mL after 48 hours). SARS-CoV-1 had similar stability kinetics (polypropylene from 10^3.4^ to 10^0.7^ TCID_50_/mL after 72 hours, stainless steel from 10^3.6^ to 10^0.6^ TCID_50_/mL after 48 hours). No viable virus could be measured after 4 hours on copper for HCoV-19 and 8 hours for SARS-CoV-1, or after 24 hours on cardboard for HCoV-19 and 8 hours for SARS-CoV-1 (Figure 1A).

**Figure 1.**
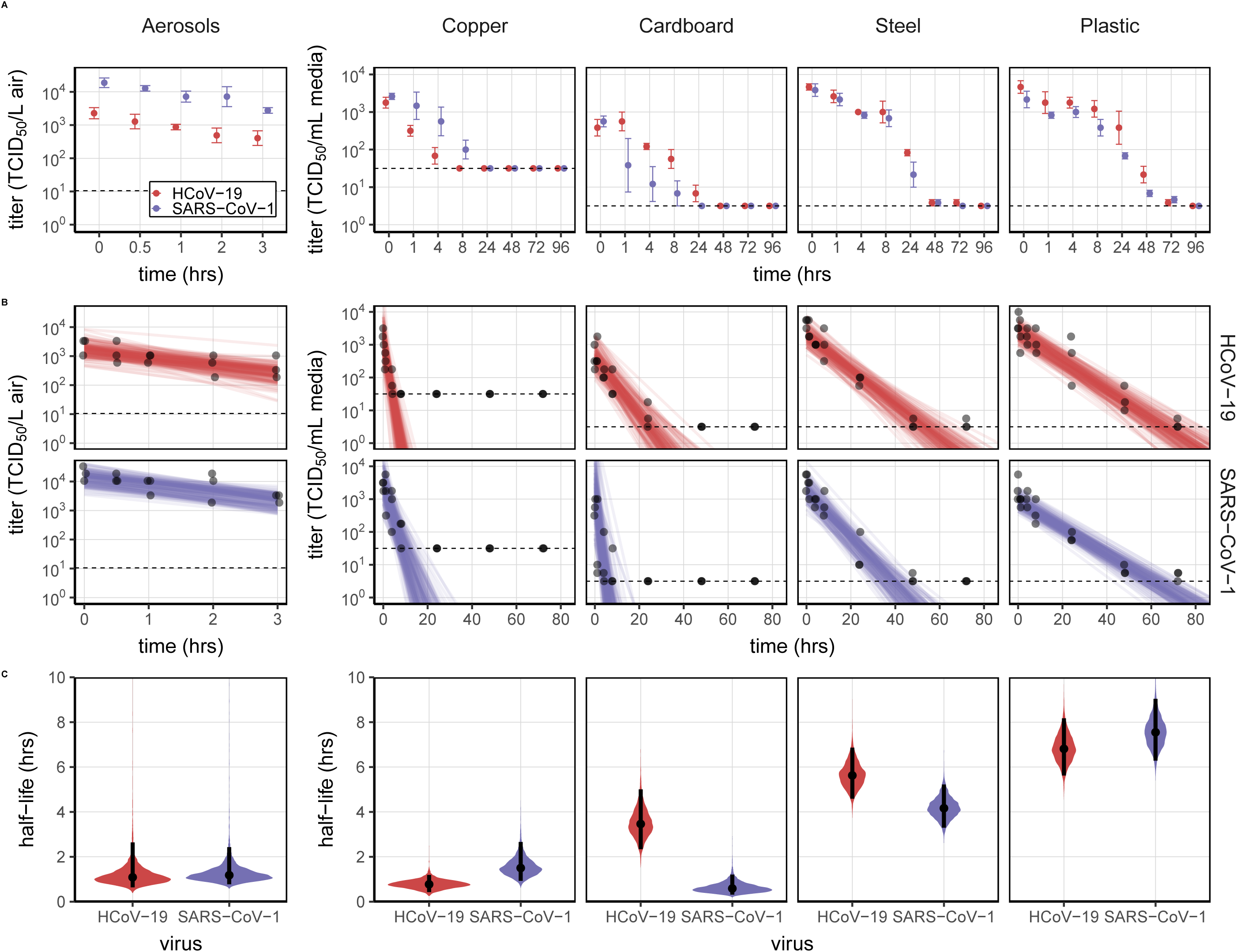
Viability of SARS-CoV-1 and HCoV-19 in aerosols and on different surfaces. A) SARS-CoV and HCoV-19 were aerosolized in a rotating drum maintained at 21-23°C and 65% RH over three hours. Viable virus titer is shown in TCID_50_/L air. For surfaces, viruses were applied on copper, cardboard, steel and plastic maintained at 21-23°C and 40% RH over seven days. Viable virus titer is shown in TCID_50_/mL collection medium. All samples were quantified by end-point titration on Vero E6 cells. Plots show the mean and standard error across three replicates. B) Regression plots showing predicted decay of virus titer over time; titer plotted on a logarithmic scale. Points show measured titers and are slightly jittered along the time axis to avoid overplotting. Lines are random draws from the joint posterior distribution of the exponential decay rate (negative of the slope) and intercept (initial virus titer), thus visualizing the range of possible decay patterns for each experimental condition. 150 lines per panel: 50 lines from each plotted replicate. C) Violin plots showing posterior distribution for half-life of viable virus based on the estimated exponential decay rates of virus titer. Dot shows the posterior median estimate and black line shows a 95% credible interval. Experimental conditions are ordered by posterior median half-life for HCoV-19. Dotted line shows Limit of Detection (LOD), 3.33 × 10^0.5^ TCID_50_/L air for aerosols, 10^0.5^ TCID_50_/mL media for plastic, steel and cardboard and 10^1.5^ TCID_50_/mL media for copper.

Both viruses exhibited exponential decay in virus titer across all experimental conditions, as indicated by linear decrease in the log_10_TCID_50_/mL over time (Figure 1B). HCoV-19 and SARS-CoV-1 exhibited similar half-lives in aerosols, with median estimates around 1.1-1.2 hours, and 95% credible intervals of [0.64, 2.64] hours for HCoV-19 and [0.78, 2.43] hours for SARS-CoV-1 (Figure 1C, Table S1). Half-lives on copper were also similar between the two viruses. On cardboard, HCoV-19 showed a considerably longer half-life than SARS-CoV-1. Both viruses showed longest viability on stainless steel and plastic: the median half-life estimate for HCoV-19 was roughly 5.6 hours on steel and 6.8 hours on plastic (Figure 1C, Table S1). Estimated differences in half-life between the two viruses were small except for on cardboard (Figure 1C, Table S1). Individual replicate data were noticeably noisier for cardboard than other surfaces (Figures S1–S5), so we advise caution in interpreting this result.

Our findings show that the stability of HCoV-19 and SARS-CoV-1 under the experimental circumstances tested is similar. This indicates that differences in the epidemiology of these viruses likely arise from other factors, including high viral loads in the upper respiratory tract and the potential for individuals infected with HCoV-19 to shed and transmit the virus while asymptomatic^3,4^. Our results indicate that aerosol and fomite transmission of HCoV-19 are plausible, as the virus can remain viable and infectious in aerosols for multiple hours and on surfaces up to days. This echoes the experience with SARS-CoV-1, where these modes of transmission were associated with nosocomial spread and superspreading events^5^, and provides guidance for pandemic mitigation measures.

## Data Availability

data is available upon request

